## Supplementary Information for "Effectiveness of mRNA-1273 bivalent (Original and Omicron BA.4/BA.5) COVID-19 vaccine in preventing hospitalizations for COVID-19, medically attended SARS-CoV-2 infections, and hospital death in the United States"

**Affiliations:**

### **Supplementary Table 1.** Comparison of baseline characteristics between individuals in the bivalent (original and Omicron BA.4/BA.5) mRNA-1273 COVID-19 vaccine cohort and the COVID-19 unvaccinated cohort.

|  | **Bivalent Vaccine Group** | **COVID-19  Unvaccinated Group^a^** | **Total** | **p value** | **Absolute Standardized Difference** |
| --- | --- | --- | --- | --- | --- |
|  | **N=290292** | **N=204655** | **N=494947** |  |  |
| Age at index date, years |  |  |  | <0.01 | 0.34 |
| mean (sd) | 58.67 (17.53) | 52.55 (18.04) | 56.14 (18.00) |  |  |
| median | 62 | 53 | 58 |  |  |
| Q1, Q3 | 46, 72 | 40, 66 | 44, 70 |  |  |
| min, max | 6, 10 | 6, 11 | 6, 11 |  |  |
| Age at index date, years, n (%) |  |  |  | <0.01 | 0.32 |
| 6-17 | 2715 (0.9%) | 2715 (1.3%) | 5430 (1.1%) |  |  |
| 18-44 | 63953 (22.0%) | 57434 (28.1%) | 121387 (24.5%) |  |  |
| 45-64 | 96293 (33.2%) | 85376 (41.7%) | 181669 (36.7%) |  |  |
| 65-74 | 73258 (25.2%) | 37473 (18.3%) | 110731 (22.4%) |  |  |
| ≥75 | 54073 (18.6%) | 21657 (10.6%) | 75730 (15.3%) |  |  |
| Sex, n (%) |  |  |  | <0.01 | 0.02 |
| Female | 157727 (54.3%) | 112732 (55.1%) | 270459 (54.6%) |  |  |
| Male | 132565 (45.7%) | 91923 (44.9%) | 224488 (45.4%) |  |  |
| Race/Ethnicity, n (%) |  |  |  | <0.01 | 0.27 |
| Non-Hispanic White | 114740 (39.5%) | 85828 (41.9%) | 200568 (40.5%) |  |  |
| Non-Hispanic Black | 23517 (8.1%) | 16662 (8.1%) | 40179 (8.1%) |  |  |
| Hispanic | 82547 (28.4%) | 67279 (32.9%) | 149826 (30.3%) |  |  |
| Non-Hispanic Asian | 50129 (17.3%) | 17459 (8.5%) | 67588 (13.7%) |  |  |
| Other/Unknown | 19359 (6.7%) | 17427 (8.5%) | 36786 (7.4%) |  |  |
| Body mass index^b^, kg/m^2^, n (%) |  |  |  | <0.01 | 0.24 |
| <18.5 | 5233 (1.8%) | 4291 (2.1%) | 9524 (1.9%) |  |  |
| 18.5 - <25 | 75876 (26.1%) | 44238 (21.6%) | 120114 (24.3%) |  |  |
| 25 - <30 | 89285 (30.8%) | 56946 (27.8%) | 146231 (29.5%) |  |  |
| 30 - <35 | 53028 (18.3%) | 37658 (18.4%) | 90686 (18.3%) |  |  |
| 35 - <40 | 24218 (8.3%) | 17156 (8.4%) | 41374 (8.4%) |  |  |
| 40 - <45 | 10083 (3.5%) | 6790 (3.3%) | 16873 (3.4%) |  |  |
| ≥45 | 6432 (2.2%) | 4104 (2.0%) | 10536 (2.1%) |  |  |
| Unknown | 26137 (9.0%) | 33472 (16.4%) | 59609 (12.0%) |  |  |
| Smoking^b^, n (%) |  |  |  | <0.01 | 0.19 |
| No | 216879 (74.7%) | 142550 (69.7%) | 359429 (72.6%) |  |  |
| Yes | 55588 (19.1%) | 38415 (18.8%) | 94003 (19.0%) |  |  |
| Unknown | 17825 (6.1%) | 23690 (11.6%) | 41515 (8.4%) |  |  |
| Charlson comorbidity score^c^, n (%) |  |  |  | <0.01 | 0.33 |
| 0 | 162456 (56.0%) | 145154 (70.9%) | 307610 (62.2%) |  |  |
| 1 | 50534 (17.4%) | 27666 (13.5%) | 78200 (15.8%) |  |  |
| ≥2 | 77302 (26.6%) | 31835 (15.6%) | 109137 (22.1%) |  |  |
| Frailty index^c^ |  |  |  | <0.01 | 0.16 |
| mean (sd) | 0.12 (0.03) | 0.12 (0.03) | 0.12 (0.03) |  |  |
| median | 0.11 | 0.11 | 0.11 |  |  |
| Q1, Q3 | 0.10, 0.14 | 0.10, 0.13 | 0.10, 0.13 |  |  |
| min, max | 0.04, 0.41 | 0.04, 0.41 | 0.04, 0.41 |  |  |
| Frailty index^c^, n (%) |  |  |  | <0.01 | 0.35 |
| Quartile 1 | 77937 (26.8%) | 45784 (22.4%) | 123721 (25.0%) |  |  |
| Quartile 2 | 55217 (19.0%) | 68551 (33.5%) | 123768 (25.0%) |  |  |
| Quartile 3 | 74608 (25.7%) | 49109 (24.0%) | 123717 (25.0%) |  |  |
| Quartile 4, most frail | 82530 (28.4%) | 41211 (20.1%) | 123741 (25.0%) |  |  |
| Chronic diseases^c^, n (%) |  |  |  |  |  |
| Kidney disease | 28145 (9.7%) | 10538 (5.1%) | 38683 (7.8%) | <0.01 | 0.17 |
| Heart disease | 14069 (4.8%) | 7477 (3.7%) | 21546 (4.4%) | <0.01 | 0.06 |
| Lung disease | 34966 (12.0%) | 18063 (8.8%) | 53029 (10.7%) | <0.01 | 0.11 |
| Liver disease | 11715 (4.0%) | 5996 (2.9%) | 17711 (3.6%) | <0.01 | 0.06 |
| Diabetes | 58781 (20.2%) | 24753 (12.1%) | 83534 (16.9%) | <0.01 | 0.22 |
| Immunocompromised status, n (%) |  |  |  | <0.01 | 0.11 |
| Yes | 12338 (4.3%) | 4788 (2.3%) | 17126 (3.5%) |  |  |
| HIV/AIDS | 1894 | 295 | 2189 |  |  |
| Leukemia, lymphoma, congenital and other immunodeficiencies, asplenia/hyposplenia | 5252 | 2299 | 7551 |  |  |
| Organ transplant | 1205 | 325 | 1530 |  |  |
| Immunosuppressant medications | 6343 | 2511 | 8854 |  |  |
| Autoimmune conditions^c^, n (%) |  |  |  | <0.01 | 0.08 |
| Yes | 12183 (4.2%) | 5650 (2.8%) | 17833 (3.6%) |  |  |
| Rheumatoid arthritis | 5206 | 2549 | 7755 |  |  |
| Inflammatory bowel disease | 2063 | 971 | 3034 |  |  |
| Psoriasis and psoriatic arthritis | 4641 | 1930 | 6571 |  |  |
| Multiple sclerosis | 614 | 357 | 971 |  |  |
| Systemic lupus erythematosus | 807 | 435 | 1242 |  |  |
| Pregnant at index date, n (%) |  |  |  | <0.01 | 0.06 |
| Yes | 1200 (0.4%) | 1759 (0.9%) | 2959 (0.6%) |  |  |
| 1st trimester | 280 | 503 | 783 |  |  |
| 2nd trimester | 462 | 610 | 1072 |  |  |
| 3rd trimester | 458 | 646 | 1104 |  |  |
| History of SARS-CoV-2 infection^d^, n (%) |  |  |  | <0.01 | 0.19 |
| Yes | 69405 (23.9%) | 66117 (32.3%) | 135522 (27.4%) |  |  |
| ≤180 days | 31592 | 13526 | 45118 |  |  |
| 181-365 days | 20310 | 25857 | 46167 |  |  |
| >365 days | 17503 | 26734 | 44237 |  |  |
| History of SARS-CoV-2 molecular test^d^, n (%) | 194122 (66.9%) | 118191 (57.8%) | 312313 (63.1%) | <0.01 | 0.19 |
| Number of outpatient and virtual visits^c^, n (%) |  |  |  | <0.01 | 0.56 |
| 0 | 12737 (4.4%) | 28594 (14.0%) | 41331 (8.4%) |  |  |
| 1-4 | 68261 (23.5%) | 78391 (38.3%) | 146652 (29.6%) |  |  |
| 5-10 | 87609 (30.2%) | 50454 (24.7%) | 138063 (27.9%) |  |  |
| ≥11 | 121685 (41.9%) | 47216 (23.1%) | 168901 (34.1%) |  |  |
| Number of Emergency Department visits^c^, n (%) |  |  |  | <0.01 | 0.08 |
| 0 | 244234 (84.1%) | 166007 (81.1%) | 410241 (82.9%) |  |  |
| 1 | 32951 (11.4%) | 26830 (13.1%) | 59781 (12.1%) |  |  |
| ≥2 | 13107 (4.5%) | 11818 (5.8%) | 24925 (5.0%) |  |  |
| Number of hospitalizations^c^, n (%) |  |  |  | <0.01 | 0.05 |
| 0 | 276000 (95.1%) | 192324 (94.0%) | 468324 (94.6%) |  |  |
| 1 | 11132 (3.8%) | 9575 (4.7%) | 20707 (4.2%) |  |  |
| ≥2 | 3160 (1.1%) | 2756 (1.3%) | 5916 (1.2%) |  |  |
| Preventive care^c^, n (%) | 251979 (86.8%) | 111295 (54.4%) | 363274 (73.4%) | <0.01 | 0.76 |
| Medicaid, n (%) | 17571 (6.1%) | 23186 (11.3%) | 40757 (8.2%) | <0.01 | 0.19 |
| Neighborhood median household income, n (%) |  |  |  | <0.01 | 0.17 |
| < $40,000 | 8206 (2.8%) | 8353 (4.1%) | 16559 (3.3%) |  |  |
| $40,000-$59,999 | 44119 (15.2%) | 36800 (18.0%) | 80919 (16.3%) |  |  |
| $60,000-$79,999 | 62206 (21.4%) | 50238 (24.5%) | 112444 (22.7%) |  |  |
| ≥$80,000 | 175589 (60.5%) | 108468 (53.0%) | 284057 (57.4%) |  |  |
| Unknown | 172 (0.1%) | 796 (0.4%) | 968 (0.2%) |  |  |
| Concomitant vaccination^e^, n (%) | 50729 (17.5%) | N/A | N/A | N/A | N/A |
| Antiviral therapy^f^, n (%) |  |  |  | <0.01 | 0.09 |
| Yes | 4094 (1.4%) | 1126 (0.6%) | 5220 (1.1%) |  |  |
| Nirmatrelvir/ritonavir | 4058 | 1113 | 5171 |  |  |
| Molnupiravir | 36 | 10 | 46 |  |  |
| Remdesivir | 3 | 3 | 6 |  |  |
| Medical center area^g^, n (%) |  |  |  | <0.01 | 0.52 |
| Month of index date, n (%) |  |  |  | <0.01 | 0.03 |
| September 2022 | 45806 (15.8%) | 32853 (16.1%) | 78659 (15.9%) |  |  |
| October 2022 | 88322 (30.4%) | 59849 (29.2%) | 148171 (29.9%) |  |  |
| November 2022 | 87272 (30.1%) | 62030 (30.3%) | 149302 (30.2%) |  |  |
| December 2022 | 68892 (23.7%) | 49923 (24.4%) | 118815 (24.0%) |  |  |

Min, minimum; max, maximum; N/A, not applicable; Q, quartile; sd, standard deviation

^a^Not all bivalent-vaccinated individuals were matched to an COVID-19 unvaccinated comparator. Unmatched bivalent-vaccinated individuals were kept in the analysis.

^b^Defined in the two years prior to index date.

^c^Defined in the one year prior to index date.

^d^Defined based on all available medical records from March 1, 2020 to index date.

^e^Among subjects with concomitant vaccines received with the bivalent mRNA-1273 vaccine: influenza vaccine (89.6%), shingles vaccine (9.1%), pneumococcal vaccine (2.9%), Tdap (2.6%), and other vaccine (1.3%).

^f^Defined during follow-up.

^g^Frequency and percent for the 19 medical center areas not shown.

### **Supplementary Table 2.** Incidence rate, hazard ratio, and relative effectiveness of the bivalent (original and Omicron BA.4/BA.5) mRNA-1273 COVID-19 vaccine in preventing hospitalization for COVID-19, overall and by subgroups (≥2 monovalent mRNA vaccine group as comparator).

|  | **Bivalent vaccine group** | | | | **≥2 monovalent mRNA vaccine group** | | | | **Hazard Ratio (95% CI)** | | **rVE (95% CI)** | |
| --- | --- | --- | --- | --- | --- | --- | --- | --- | --- | --- | --- | --- |
| **Hospitalization for COVID-19** | **N** | **Number of cases** | **Number of person years** | **Incidence per 1000 person-years  (95% CI)** | **N** | **Number of cases** | **Number of person years** | **Incidence per 1000 person-years (95% CI)** | **Unadjusted** | **Adjusted^a^** | **Unadjusted** | **Adjusted^a^** |
| Overall | 290292 | 160 | 59235.05 | 2.70  (2.31-3.15) | 580584 | 646 | 76386.05 | 8.46  (7.83-9.14) | 0.34  (0.28-0.40) | 0.30  (0.25-0.36) | 66.3  (59.9-71.7) | 70.3  (64.0-75.4) |
| Age at index date, years^b^ |  |  |  |  |  |  |  |  |  |  |  |  |
| 6-17 | 2715 | 0 | 418 | N/A | 5430 | 0 | 698.51 | N/A | N/A | N/A | N/A | N/A |
| 18-44 | 63953 | 2 | 13001.62 | 0.15  (0.04-0.62) | 127906 | 10 | 20439.67 | 0.49  (0.26-0.91) | 0.31  (0.07-1.44) | 0.22  (0.04-1.15) | 68.6  (-30.5-93.1) | 78.4  (-12.7-95.9) |
| 45-64 | 96293 | 19 | 19008.44 | 1.00  (0.64-1.57) | 192586 | 49 | 26447.6 | 1.85  (1.40-2.45) | 0.56  (0.33-0.95) | 0.44  (0.25-0.77) | 44.5  (5.4-67.4) | 56.2  (22.5-75.2) |
| 65-74 | 73258 | 41 | 15402.42 | 2.66  (1.96-3.62) | 146516 | 148 | 16799.18 | 8.81  (7.50-10.35) | 0.32  (0.23-0.46) | 0.31  (0.21-0.45) | 67.7  (54.1-77.2) | 69.3  (55.2-79.0) |
| ≥75 | 54073 | 98 | 11404.56 | 8.59  (7.05-10.47) | 108146 | 439 | 12001.09 | 36.58  (33.31-40.17) | 0.24  (0.19-0.30) | 0.29  (0.22-0.36) | 75.7  (69.7-80.6) | 71.4  (63.6-77.6) |
| Sex |  |  |  |  |  |  |  |  |  |  |  |  |
| Female | 157727 | 81 | 32124.22 | 2.52  (2.03-3.13) | 315454 | 303 | 41338.32 | 7.33  (6.55-8.20) | 0.36  (0.28-0.46) | 0.34  (0.26-0.44) | 64.0  (53.9-71.9) | 66.5  (56.0-74.5) |
| Male | 132565 | 79 | 27110.83 | 2.91  (2.34-3.63) | 265130 | 343 | 35047.72 | 9.79  (8.80-10.88) | 0.32  (0.25-0.41) | 0.27  (0.20-0.35) | 68.3  (59.5-75.2) | 73.4  (65.3-79.7) |
| Race/Ethnicity |  |  |  |  |  |  |  |  |  |  |  |  |
| Non-Hispanic White | 114740 | 77 | 24473.05 | 3.15  (2.52-3.93) | 229480 | 309 | 29162.19 | 10.60  (9.48-11.85) | 0.32  (0.25-0.41) | 0.30  (0.23-0.40) | 68.2  (59.1-75.3) | 70.0  (60.2-77.4) |
| Non-Hispanic Black | 23517 | 17 | 4520.19 | 3.76  (2.34-6.05) | 47034 | 71 | 5720.44 | 12.41  (9.84-15.66) | 0.31  (0.18-0.53) | 0.26  (0.15-0.47) | 68.8  (46.7-81.7) | 73.6  (53.0-85.1) |
| Hispanic | 82547 | 44 | 15843.06 | 2.78  (2.07-3.73) | 165094 | 166 | 23173.71 | 7.16  (6.15-8.34) | 0.40  (0.29-0.56) | 0.36  (0.25-0.52) | 59.8  (43.9-71.2) | 63.8  (48.4-74.6) |
| Non-Hispanic Asian | 50129 | 21 | 10450.58 | 2.01  (1.31-3.08) | 100258 | 84 | 12661.84 | 6.63  (5.36-8.22) | 0.31  (0.19-0.51) | 0.24  (0.14-0.40) | 68.6  (49.0-80.6) | 76.3  (59.9-86.0) |
| Immunocompromised status |  |  |  |  |  |  |  |  |  |  |  |  |
| Yes^c^ | 12338 | 30 | 2553.68 | 11.75  (8.21-16.80) | 19991 | 83 | 2289.59 | 36.25  (29.23-44.95) | 0.33  (0.22-0.51) | 0.35  (0.22-0.56) | 66.7  (49.3-78.2) | 64.7  (44.0-77.7) |
| No | 277954 | 130 | 56681.38 | 2.29  (1.93-2.72) | 560593 | 563 | 74096.45 | 7.60  (7.00-8.25) | 0.32  (0.26-0.39) | 0.29  (0.23-0.35) | 68.1  (61.3-73.6) | 71.3  (64.5-76.7) |
| History of SARS-CoV-2 infection |  |  |  |  |  |  |  |  |  |  |  |  |
| Yes^d^ | 69405 | 21 | 13177.97 | 1.59  (1.04-2.44) | 155360 | 82 | 21153.77 | 3.88  (3.12-4.81) | 0.42  (0.26-0.68) | 0.39  (0.23-0.66) | 58.1  (32.1-74.1) | 60.7  (33.8-76.7) |
| No | 220887 | 139 | 46057.08 | 3.02  (2.56-3.56) | 425224 | 564 | 55232.27 | 10.21  (9.40-11.09) | 0.31  (0.26-0.38) | 0.29  (0.24-0.35) | 68.5  (62.1-73.9) | 71.1  (64.6-76.5) |
| Months of follow-up |  |  |  |  |  |  |  |  |  |  |  |  |
| 0-<1 month | 290292 | 56 | 23761.66 | 2.36  (1.81-3.06) | 580584 | 388 | 39288.55 | 9.88  (8.94-10.91) | 0.24  (0.18-0.31) | 0.25  (0.19-0.34) | 76.2  (68.5-82.0) | 74.9  (66.4-81.3) |
| 1-<2 months | 265962 | 45 | 18967.95 | 2.37  (1.77-3.18) | 378308 | 197 | 24055.89 | 8.19  (7.12-9.42) | 0.29  (0.21-0.40) | 0.24  (0.17-0.35) | 70.9  (59.7-78.9) | 75.8  (65.3-83.2) |
| 2-<3 months | 190879 | 46 | 11685.44 | 3.94  (2.95-5.26) | 211800 | 52 | 10801.58 | 4.81  (3.67-6.32) | 0.84  (0.56-1.24) | 0.41  (0.25-0.67) | 16.5  (-19.6-43.9) | 58.9  (33.5-74.6) |
| ≥3 months | 92435 | 13 | 4413.99 | 2.95  (1.71-5.07) | 62428 | 9 | 2138.48 | 4.21  (2.19-8.09) | 0.82  (0.35-1.92) | 0.20  (0.07-0.57) | 18.2  (-48.0-65.2) | 79.6  (43.2-92.7) |
| Number of monovalent vaccines prior to index date^e^ | |  |  |  |  |  |  |  |  |  |  |  |
| 2 doses | 11493 | 3 | 2203.2 | 1.36  (0.44-4.22) | 135437 | 149 | 22354.78 | 6.67  (5.68-7.83) | 0.22  (0.07-0.69) | 0.23  (0.07-0.72) | 78.0  (31.0-93.0) | 77.3  (27.6-92.9) |
| 3 doses | 144052 | 48 | 28340.77 | 1.69  (1.28-2.25) | 287077 | 311 | 39692.37 | 7.84  (7.01-8.76) | 0.23  (0.17-0.31) | 0.30  (0.22-0.40) | 77.2  (69.0-83.2) | 70.4  (59.6-78.3) |
| ≥4 doses | 134747 | 109 | 28691.09 | 3.80  (3.15-4.58) | 158070 | 186 | 14338.9 | 12.97  (11.24-14.98) | 0.29  (0.22-0.37) | 0.28  (0.22-0.36) | 71.3  (63.3-77.5) | 71.7  (63.7-78.0) |
| Time between latest monovalent vaccine and index date^f^ | |  |  |  |  |  |  |  |  |  |  |  |
| ≤180 days | 83060 | 83 | 19923.17 | 4.17  (3.36-5.17) | 134392 | 142 | 13192.87 | 10.76  (9.13-12.69) | 0.40  (0.30-0.52) | 0.36  (0.27-0.48) | 60.3  (47.6-70.0) | 64.0  (51.7-73.2) |
| 181-365 days | 165276 | 64 | 33439.32 | 1.91  (1.50-2.45) | 274298 | 281 | 37735.09 | 7.45  (6.62-8.37) | 0.27  (0.20-0.35) | 0.22  (0.17-0.29) | 73.3  (65.0-79.7) | 78.0  (71.1-83.3) |
| >365 days | 41956 | 13 | 5872.56 | 2.21  (1.29-3.81) | 171894 | 223 | 25458.09 | 8.76  (7.68-9.99) | 0.25  (0.14-0.43) | 0.26  (0.15-0.46) | 75.5  (57.1-86.0) | 73.6  (53.7-85.0) |

CI, confidence interval; rVE, relative vaccine effectiveness

^a^Adjusted for covariates age group, sex, race/ethnicity, index date (in months), history of SARS-CoV-2 infection, number of outpatient and virtual visits, preventive care, number of monovalent vaccines prior to index date, time between latest monovalent vaccine and index date, and antiviral therapy. Medical center area removed from adjustment set due to lack of model convergence.

^b^Adjusted for continuous age (in years) in addition to covariates above.

^c^Adjusted for immunocompromising sub-conditions in addition to covariates above.

^d^Adjusted for time since prior SARS-CoV-2 infection in addition to covariates above.

^e^Time between latest monovalent vaccine and index date removed from adjustment set due to lack of model convergence.

^f^Number of monovalent vaccines prior to index date removed from adjustment set due to lack of model convergence.

When the hazard ratio or its 95% CI was >1, the rVE or its 95% CI was transformed as ([1/hazard ratio] – 1) × 100

### **Supplementary Table 3.** Incidence rate, hazard ratio, and effectiveness of the bivalent (original and Omicron BA.4/BA.5) mRNA-1273 COVID-19 vaccine in preventing hospitalization for COVID-19, overall and by subgroups (COVID-19 unvaccinated group as comparator).

|  | **Bivalent vaccine group** | | | | **COVID-19 unvaccinated group** | | | | **Hazard Ratio (95% CI)** | | **VE (95% CI)** | |
| --- | --- | --- | --- | --- | --- | --- | --- | --- | --- | --- | --- | --- |
| **Hospitalization for COVID-19** | **N** | **Number of cases** | **Number of person years** | **Incidence per 1000 person-years  (95% CI)** | **N** | **Number of cases** | **Number of person years** | **Incidence per 1000 person-years (95% CI)** | **Unadjusted** | **Adjusted^a^** | **Unadjusted** | **Adjusted^a^** |
| Overall | 290292 | 160 | 59235.05 | 2.70  (2.31-3.15) | 204655 | 341 | 39937.04 | 8.54  (7.68-9.49) | 0.32  (0.26-0.38) | 0.17  (0.14-0.21) | 68.4  (61.9-73.8) | 82.8  (78.8-86.0) |
| Age at index date, years^b^ |  |  |  |  |  |  |  |  |  |  |  |  |
| 6-17 | 2715 | 0 | 418 | N/A | 2715 | 0 | 411.76 | N/A | N/A | N/A | N/A | N/A |
| 18-44^c,d^ | 63953 | 2 | 13001.62 | 0.15  (0.04-0.62) | 57434 | 5 | 11004.34 | 0.45  (0.19-1.09) | 0.34  (0.07-1.74) | 0.13  (0.02-0.89) | 66.2  (-42.7-93.4) | 87.5  (10.5-98.2) |
| 45-64 | 96293 | 19 | 19008.44 | 1.00  (0.64-1.57) | 85376 | 43 | 16201.03 | 2.65  (1.97-3.58) | 0.38  (0.22-0.65) | 0.22  (0.12-0.39) | 62.2  (35.1-77.9) | 78.4  (61.2-87.9) |
| 65-74 | 73258 | 41 | 15402.42 | 2.66  (1.96-3.62) | 37473 | 91 | 7774.11 | 11.71  (9.53-14.38) | 0.23  (0.16-0.33) | 0.18  (0.12-0.27) | 77.3  (67.2-84.3) | 82.1  (73.2-88.0) |
| ≥75 | 54073 | 98 | 11404.56 | 8.59  (7.05-10.47) | 21657 | 202 | 4545.79 | 44.44  (38.71-51.01) | 0.19  (0.15-0.25) | 0.16  (0.12-0.21) | 80.7  (75.4-84.8) | 84.3  (79.4-88.1) |
| Sex |  |  |  |  |  |  |  |  |  |  |  |  |
| Female | 157727 | 81 | 32124.22 | 2.52  (2.03-3.13) | 112732 | 192 | 22033.95 | 8.71  (7.56-10.04) | 0.29  (0.22-0.37) | 0.18  (0.14-0.24) | 71. (62.6-77.7) | 81.8  (75.8-86.3) |
| Male | 132565 | 79 | 27110.83 | 2.91  (2.34-3.63) | 91923 | 149 | 17903.09 | 8.32  (7.09-9.77) | 0.35  (0.27-0.46) | 0.16  (0.12-0.22) | 65.0  (54.1-73.4) | 83.7  (77.7-88.0) |
| Race/Ethnicity |  |  |  |  |  |  |  |  |  |  |  |  |
| Non-Hispanic White | 114740 | 77 | 24473.05 | 3.15  (2.52-3.93) | 85828 | 183 | 17737.23 | 10.32  (8.93-11.93) | 0.30  (0.23-0.40) | 0.16  (0.12-0.21) | 69.5  (60.3-76.7) | 84.0  (78.5-88.1) |
| Non-Hispanic Black | 23517 | 17 | 4520.19 | 3.76  (2.34-6.05) | 16662 | 44 | 3043.7 | 14.46  (10.76-19.43) | 0.26  (0.15-0.45) | 0.14  (0.08-0.26) | 74.0  (54.5-85.2) | 86.1  (74.4-92.5) |
| Hispanic | 82547 | 44 | 15843.06 | 2.78  (2.07-3.73) | 67279 | 82 | 12510.61 | 6.55  (5.28-8.14) | 0.42  (0.29-0.61) | 0.23  (0.15-0.34) | 57.6  (38.9-70.6) | 77.0  (65.5-84.6) |
| Non-Hispanic Asian | 50129 | 21 | 10450.58 | 2.01  (1.31-3.08) | 17459 | 27 | 3260 | 8.28  (5.68-12.08) | 0.24  (0.14-0.42) | 0.11  (0.06-0.21) | 76.1  (57.7-86.5) | 89.2  (78.9-94.5) |
| Immunocompromised status |  |  |  |  |  |  |  |  |  |  |  |  |
| Yes^c,e^ | 12338 | 30 | 2553.68 | 11.75  (8.21-16.80) | 4788 | 25 | 952.23 | 26.25  (17.74-38.85) | 0.45  (0.26-0.76) | 0.28  (0.16-0.51) | 55.3  (24.0-73.7) | 71.8  (48.8-84.5) |
| No | 277954 | 130 | 56681.38 | 2.29  (1.93-2.72) | 199867 | 316 | 38984.81 | 8.11  (7.26-9.05) | 0.28  (0.23-0.35) | 0.16  (0.13-0.20) | 71.  (65.4-77.0) | 84.1  (80.1-87.4) |
| History of SARS-CoV-2 infection |  |  |  |  |  |  |  |  |  |  |  |  |
| Yes^f^ | 69405 | 21 | 13177.97 | 1.59  (1.04-2.44) | 66117 | 58 | 12742.28 | 4.55  (3.52-5.89) | 0.35  (0.21-0.58) | 0.32  (0.18-0.55) | 64.  (42.1-78.7) | 68.3  (45.4-81.6) |
| No | 220887 | 139 | 46057.08 | 3.02  (2.56-3.56) | 138538 | 283 | 27194.75 | 10.41  (9.26-11.69) | 0.29  (0.24-0.35) | 0.16  (0.13-0.20) | 71.0  (64.5-76.4) | 84.3  (80.3-87.5) |
| Months of follow-up |  |  |  |  |  |  |  |  |  |  |  |  |
| 0-<1 month | 290292 | 56 | 23761.66 | 2.36  (1.81-3.06) | 204655 | 124 | 16474.69 | 7.53  (6.31-8.98) | 0.31  (0.23-0.43) | 0.15  (0.11-0.22) | 68.7  (57.1-77.2) | 84.8  (78.5-89.3) |
| 1-<2 months | 265962 | 45 | 18967.95 | 2.37  (1.77-3.18) | 181530 | 115 | 12788.96 | 8.99  (7.49-10.80) | 0.26  (0.19-0.37) | 0.14  (0.10-0.21) | 73.6  (62.7-81.3) | 85.6  (79.0-90.2%) |
| 2-<3 months | 190879 | 46 | 11685.44 | 3.94  (2.95-5.26) | 126732 | 84 | 7627.96 | 11.01  (8.89-13.64) | 0.36  (0.25-0.51) | 0.23  (0.15-0.35) | 64.2  (48.7-75.0) | 76.8  (65.0-84.6) |
| ≥3 months | 92435 | 13 | 4413.99 | 2.95  (1.71-5.07) | 58823 | 18 | 2797.68 | 6.43  (4.05-10.21) | 0.46  (0.23-0.94) | 0.25  (0.11-0.56) | 54.1  (6.2-77.5) | 75.5  (43.8-89.3) |

CI, confidence interval; VE, vaccine effectiveness

^a^Adjusted for covariates age group, sex, race/ethnicity, index date (in months), body mass index, smoking, Charlson comorbidity score, frailty index, kidney disease, lung disease, diabetes, immunocompromised status, history of SARS-CoV-2 infection, history of SARS-CoV-2 molecular test, number of outpatient and virtual visits, preventive care, Medicaid, and antiviral therapy. Neighborhood median household income and medical center area removed from adjustment set due to lack of model convergence.

^b^Adjusted for continuous age (in years) in addition to covariates above.

^c^Smoking removed from adjustment set due to lack of model convergence.

^d^Kidney disease, lung disease, and immunocompromised status removed from adjustment set due to lack of model convergence.

^e^Adjusted for immunocompromising sub-conditions in addition to covariates above.

^f^Adjusted for time since prior SARS-CoV-2 infection in addition to covariates above.

When the hazard ratio or its 95% CI was >1, the VE or its 95% CI was transformed as ([1/hazard ratio] – 1) × 100.

### **Supplementary Table 4.** Incidence rate, hazard ratio, and relative effectiveness of the bivalent (original and Omicron BA.4/BA.5) mRNA-1273 COVID-19 vaccine in preventing medically attended SARS-CoV-2 infection and COVID-19 hospital death (≥2 monovalent mRNA vaccine group as comparator).

|  | **Bivalent vaccine group** | | | | **≥2 monovalent mRNA vaccine group** | | | | **Hazard Ratio (95% CI)** | | **rVE (95% CI)** | |
| --- | --- | --- | --- | --- | --- | --- | --- | --- | --- | --- | --- | --- |
| **Outcomes** | **N** | **Number of cases** | **Number of person years** | **Incidence per 1000 person-years  (95% CI)** | **N** | **Number of cases** | **Number of person years** | **Incidence per 1000 person-years (95% CI)** | **Unadjusted** | **Adjusted^a^** | **Unadjusted** | **Adjusted^a^** |
| Medically attended SARS-CoV-2 infection |  |  |  |  |  |  |  |  |  |  |  |  |
| All care settings | 290292 | 3221 | 58867.97 | 54.72  (52.86-56.64) | 580584 | 4964 | 75886.22 | 65.41  (63.62-67.26) | 0.84  (0.80-0.88) | 0.64  (0.61-0.67) | 16.0  (12.1-19.7) | 35.9  (32.7-39.0) |
| Emergency department and urgent care | 290292 | 855 | 59149.21 | 14.45  (13.52-15.46) | 580584 | 2083 | 76214.31 | 27.33  (26.18-28.53) | 0.55  (0.51-0.60) | 0.45  (0.41-0.49) | 45.0  (40.4-49.2) | 55.0  (50.8-58.8) |
| COVID-19 hospital death^b^ | 290292 | 10 | 59252.1 | 0.17  (0.09-0.31) | 580584 | 59 | 76449.67 | 0.77  (0.60-1.00) | 0.22  (0.11-0.44) | 0.17  (0.08-0.36) | 77.7  (56.4-88.6) | 82.7  (63.7-91.7) |

CI, confidence interval; rVE, relative vaccine effectiveness

^a^Adjusted for covariates age group, sex, race/ethnicity, index date (in months), history of SARS-CoV-2 infection, number of outpatient and virtual visits, preventive care, number of monovalent vaccines prior to index date, time between latest monovalent vaccine and index date, and medical center area.

^b^Adjusted for antiviral therapy in addition to covariates above. Medical center area removed from adjustment set due to lack of model convergence.

### **Supplementary Table 5.** Incidence rate, hazard ratio, and effectiveness of the bivalent (original and Omicron BA.4/BA.5) mRNA-1273 COVID-19 vaccine in preventing medically attended SARS-CoV-2 infection and COVID-19 hospital death (COVID-19 unvaccinated group as comparator).

|  | **Bivalent vaccine group** | | | | **COVID-19 unvaccinated group** | | | | **Hazard Ratio (95% CI)** | | **VE (95% CI)** | |
| --- | --- | --- | --- | --- | --- | --- | --- | --- | --- | --- | --- | --- |
| **Outcomes** | **N** | **Number of cases** | **Number of person years** | **Incidence per 1000 person-years  (95% CI)** | **N** | **Number of cases** | **Number of person years** | **Incidence per 1000 person-years (95% CI)** | **Unadjusted** | **Adjusted^a^** | **Unadjusted** | **Adjusted^a^** |
| Medically attended SARS-CoV-2 infection |  |  |  |  |  |  |  |  |  |  |  |  |
| All care settings | 290292 | 3221 | 58867.97 | 54.72  (52.86-56.64) | 204655 | 1578 | 39779.99 | 39.67  (37.76-41.67) | 1.38  (1.30-1.46) | 0.89  (0.83-0.96) | -27.3  (-31.6--22.8) | 10.7  (4.4-16.6) |
| Emergency department and urgent care | 290292 | 855 | 59149.21 | 14.45  (13.52-15.46) | 204655 | 815 | 39872.1 | 20.44  (19.08-21.89) | 0.71  (0.64-0.78) | 0.45  (0.40-0.50) | 29.3  (22.1-35.7) | 55.4  (50.3-60.1) |
| COVID-19 hospital death^b^ | 290292 | 10 | 59252.1 | 0.17  (0.09-0.31) | 204655 | 35 | 39972.07 | 0.88  (0.63-1.22) | 0.19  (0.09-0.39) | 0.10  (0.05-0.22) | 80.9  (61.5-90.5) | 89.7  (77.7-95.2) |

CI, confidence interval; VE, vaccine effectiveness

^a^Adjusted for covariates age group, sex, race/ethnicity, index date (in months), body mass index, smoking, Charlson comorbidity score, frailty index, kidney disease, lung disease, diabetes, immunocompromised status, history of SARS-CoV-2 infection, history of SARS-CoV-2 molecular test, number of outpatient and virtual visits, preventive care, Medicaid, neighborhood median household income, and medical center area.

^b^Adjusted for antiviral therapy in addition to covariates above. Neighborhood median household income and medical center area removed from adjustment set due to lack of model convergence.

When the hazard ratio or its 95% CI was >1, the VE or its 95% CI was transformed as ([1/hazard ratio] – 1) × 100.
